# Low blood pressure phenotype underpins the tendency to reflex syncope

**DOI:** 10.1101/2020.11.29.20240465

**Authors:** Michele Brignole, Giulia Rivasi, Richard Sutton, Rose Anne Kenny, Carlos A Morillo, Robert Sheldon, Satish R Raj, Andrea Ungar, Raffaello Furlan, Gert van Dijk, Mohamed Hamdan, Viktor Hamrefors, Gunnar Engström, Chloe Park, Davide Soranna, Antonella Zambon, Gianfranco Parati, Artur Fedorowski

**Author notes:** Corresponding author: Michele Brignole, IRCCS Istituto Auxologico, via Magnasco 2, 20149 Milan, Italy. Co-senior Authors (Gianfranco Parati, Artur Fedorowski).

## Abstract

**BACKGROUND:** We hypothesized that cardiovascular physiology differs in reflex syncope patients compared with general population, predisposing such subjects to vasovagal reflex.

**METHODS:** In this multicohort cross-sectional study, we compared resting systolic blood pressure (SBP), diastolic blood pressure (DBP), pulse pressure (PP) and heart rate (HR), collected from 6 community-based cohort studies (64,968 observations) with those from 6 databases of reflex syncope patients (6516 observations), subdivided by age decades and sex.

**RESULTS:** Overall, in males with reflex syncope, SBP (−3.4 mmHg) and PP (−9.2 mmHg) were lower and DBP (+2.8 mmHg) and HR (+5.1 bpm) were higher than in the general population; the difference in SBP was higher at ages >60 years. In females, PP (−6.0 mmHg) was lower and DBP (+4.7 mmHg) and HR (+4.5 bpm) were higher than in the general population; differences in SBP were less pronounced, becoming evident only above 60 years. Compared with males, SBP in females exhibited slower increase until age 40, and then demonstrated steeper increase that continued throughout life.

**CONCLUSION:** The patients prone to reflex syncope demonstrate a different resting cardiovascular hemodynamic profile characterized by reduced venous return and stroke volume, evidenced by lower SBP and PP, and compensatory increase in HR and vascular resistance, the latter expressed by elevated DBP. The data presented here contribute to our understanding why some subjects with similar demographic characteristics develop reflex syncope and others not.

## Introduction

The 2018 ESC guidelines on syncope (1) introduced the concept of “low blood pressure (BP) phenotype” as a mix of clinical and investigational findings that identify those patients in whom a low BP plays a role in causing reflex (neurally-mediated) syncope. For example, the patients with persistent low BP (2-4) and those who show a hypotensive susceptibility on tilt testing (5) seem to satisfy the criteria for low BP phenotype. Being only a concept, the guidelines were unable to define more precisely low BP phenotype. Supine resting BP values have gained little importance in the literature of reflex syncope. Several trials even omitted reporting supine BP values among the baseline clinical characteristics. Among those trials that reported baseline BP values, a comparison with the general population was hampered by the fact that BP is highly age- and sex-dependent which makes the population of such trials underpowered for these purposes.

On the assumption that cardiovascular physiology may fundamentally differ in patients with reflex syncope compared with subjects from a general population, an age- and sex-specific analysis of existing data could offer new insights, improve our understanding of the mechanism of reflex syncope and help to characterize the low BP phenotype.

## Method

We compared resting BP and heart rate (HR) measures collected from 6 databases of patients affected by certain or likely reflex syncope (6516 observations) (6-13) with those collected in 6 community-based cohort studies of general population (64,968 observations)(14-19). Both syncope and general population studies were undertaken in the years 1998-2019 and were analyzed between December 2019 and April 2020.

The principal investigators of each included study were asked to calculate the average and the standard deviation of the baseline values of BP and HR subdivided by age decade and sex from his/her respective database unless the data were already available in the literature. In addition to systolic BP (SBP), diastolic BP (DBP) and HR, the variables of interest were pulse pressure (PP) and mean arterial pressure (MAP). These indices were calculated using the following formulae: PP=SBP-DBP and MAP= DBP+(1/3)PP.

The diagnosis of definite or likely reflex (neurally-mediated) syncope was made according to presence of the clinical features of reflex syncope, the exclusion of competing diagnoses (especially orthostatic hypotension) and, eventually, confirmed by cardiovascular autonomic investigations. The patients had syncope of sufficient severity to require assessment in a specialistic syncope clinic. All included studies have been approved by ethics committess of their respective study sites.

### Statistical analysis

In absence of previous data from the literature we assumed, as biologically valuable, a minimum mean difference of 2 mmHg in SBP between the syncope group and the general population group. We calculated that 250 patients with syncope and 2500 subjects from the general population for each age decade would have a power to detect a difference of 2 mmHg with a two sided alpha error of 0.10 and a beta error of 0,20. Thus, the minimum size of an age decade group was set to 2000, and for general population to 20,000.

Continuous variables were expressed as means ± SD and categorical data as frequencies and proportions. In order to evaluate the relationship of population type (general population and syncope), gender, age decade with BP and HR, we employed the Generalized Estimating Equation (GEE) models which take into account the clustering effect due to correlation among estimates within a study. Moreover, we weighted each available mean estimate for the corresponding sample size. The model included four groups of patients according to the gender and population type (syncope or general population): i) male and syncope (MSyn); ii) male and general population (MGP); iii) female and syncope (FSyn); iv) female and general population (FGP). Age decade was included as a continuous variable. To take into account the potential modifying effect of age on the association among groups and BP/HR levels, we included an interaction term between age and groups. The group-specific least square means (and their 95% confidence interval [95% CI]), adjusted for age decade were estimated. The following contrasts of least square means were performed: i) MSyn vs MGP, ii) FSyn vs FGP to underline differences among population types in males and females. Moreover, we considered a further 3 comparisons; the first two (separately for gender) related to differences in trend of BP/HR levels through age decades among syncope and general population groups and the third related only to syncope patients for differences in trend of BP/HR levels through age decades among males and females.

A sensitivity analysis was performed considering only the 3 cross-sectional consecutive series (intervention trials excluded), in order to verify the independence of findings from the potential selection biases of trials.

Finally, we evaluated the potential effect of hypertension prevalence as a confounder veryfing if the relationship between age and prevalence of hypertension changes as a function of population type. We modelled the prevalence of hypertension, available only at study level, as a function of average age and population type as well as their interaction.

All analyses were performed using the Statistical Analysis System Software (version 9.4; SAS Institute, Cary, NC). Statistical significance was set at the 0.05 level. All p values were 2-sided.

## Results

The features of the databases of patients affected by certain or likely reflex syncope and of the community-based cohort studies of general population are described in detail in the *eTable 1* and *eTable 2*. There were 44% and 45% males respectively with ages spanning from the 2^nd^ to the 9^th^ decade of life. Antihypertensive therapy rate was correlated with the respective average ages of the studies (p=0.03) and was balanced between syncope and general population cohorts without potential confounding effect, p=0.89 (*eTable 1* and *eFigure 1*).

The comparison of SBP, DBP and HR per age decade and sex between syncope patients and subjects from the general population is displayed in the **Table 1**. Overall, in males with reflex syncope, SBP (−3.4 mmHg) and PP (−9.2 mmHg) were lower and DBP (+2.8 mmHg) and HR (+5.1 bpm) were higher than in the general population; the difference in SBP was higher at ages >60 years **(Figure 1)**. In females, PP (−6.0 mmHg) was lower and DBP (+4.7 mmHg) and HR (+4.5 bpm) were higher than in the general population; differences in SBP were less pronounced, becoming evident only above 60 years **(Figure 1)**. In contrast, there was no decrease in MAP in syncope patients compared with the general population either in males (93.0 mmHg versus 92.5 mmHg respectively) and in females (91.4 mmHg versus 89.0 mmHg respectively).

**Table 1.** Overall results. Observed and least square adjusted supine BP and HR in patients with reflex syncope: comparison with general population. The group-specific least square means (and their 95% confidence interval [95% CI]), adjusted for age decade were estimated

|  | Syncope |  |  |  | General population |  |  |  | P value |
| --- | --- | --- | --- | --- | --- | --- | --- | --- | --- |
|  | N° of pts | Observed mean | LS adjusted mean | 95% CI | N° of pts | Observed mean | LS adjusted mean | 95% CI |  |
| Males |  |  |  |  |  |  |  |  |  |
| Systolic blood pressure, mmHg | 2855 | 128.1 | 126.7 | 124.0 - 129.3 | 29424 | 131.5 | 131.8 | 129.3 - 134.2 | 0.006 |
| Diastolic blood pressure, mmHg | 2426 | 77.7 | 77.4 | 75.0 - 79.7 | 29424 | 74.9 | 75.3 | 73.9 - 76.6 | 0.14 |
| Pulse pressure, mmHg | 2425 | 48,8 | 49.0 | 44.9 - 53.1 | 29424 | 58,0 | 56.2 | 54.1 - 58.2 | 0.002 |
| Mean arterial pressure, mmHg | 2425 | 93.0 | 93.7 | 92.0 - 95.4 | 29424 | 92.5 | 93.7 | 92.4 – 95.5 | 0.81 |
| Heart rate, bpm | 2020 | 67.7 | 67.9 | 66.5 - 69.3 | 9493 | 62.6 | 62.7 | 60.4 - 64.9 | 0.0001 |
| Females |  |  |  |  |  |  |  |  |  |
| Systolic blood pressure, mmHg | 3661 | 127.2 | 125.9 | 122.1 - 129.8 | 35554 | 125.7 | 127.5 | 125.1-129.9 | 0.50 |
| Diastolic blood pressure, mmHg | 2961 | 75.7 | 74.9 | 73.9 - 75.8 | 35544 | 71.0 | 71.3 | 70.3-72.4 | 0.0001 |
| Pulse pressure, mmHg | 2960 | 50,2 | 51.3 | 46.3 - 56.2 | 35544 | 56,2 | 55.5 | 52.2-58.8 | 0.16 |
| Mean arterial pressure, mmHg | 2960 | 91.4 | 92.0 | 90.7 - 93.2 | 35544 | 89.0 | 89.8 | 89.4 – 90.2 | 0.001 |
| Heart rate, bpm | 2428 | 70.2 | 70.5 | 69.6 - 71.5 | 11576 | 65.7 | 65.0 | 62.3-67.6 | 0.0001 |

LS=least square

**Figure 1.**
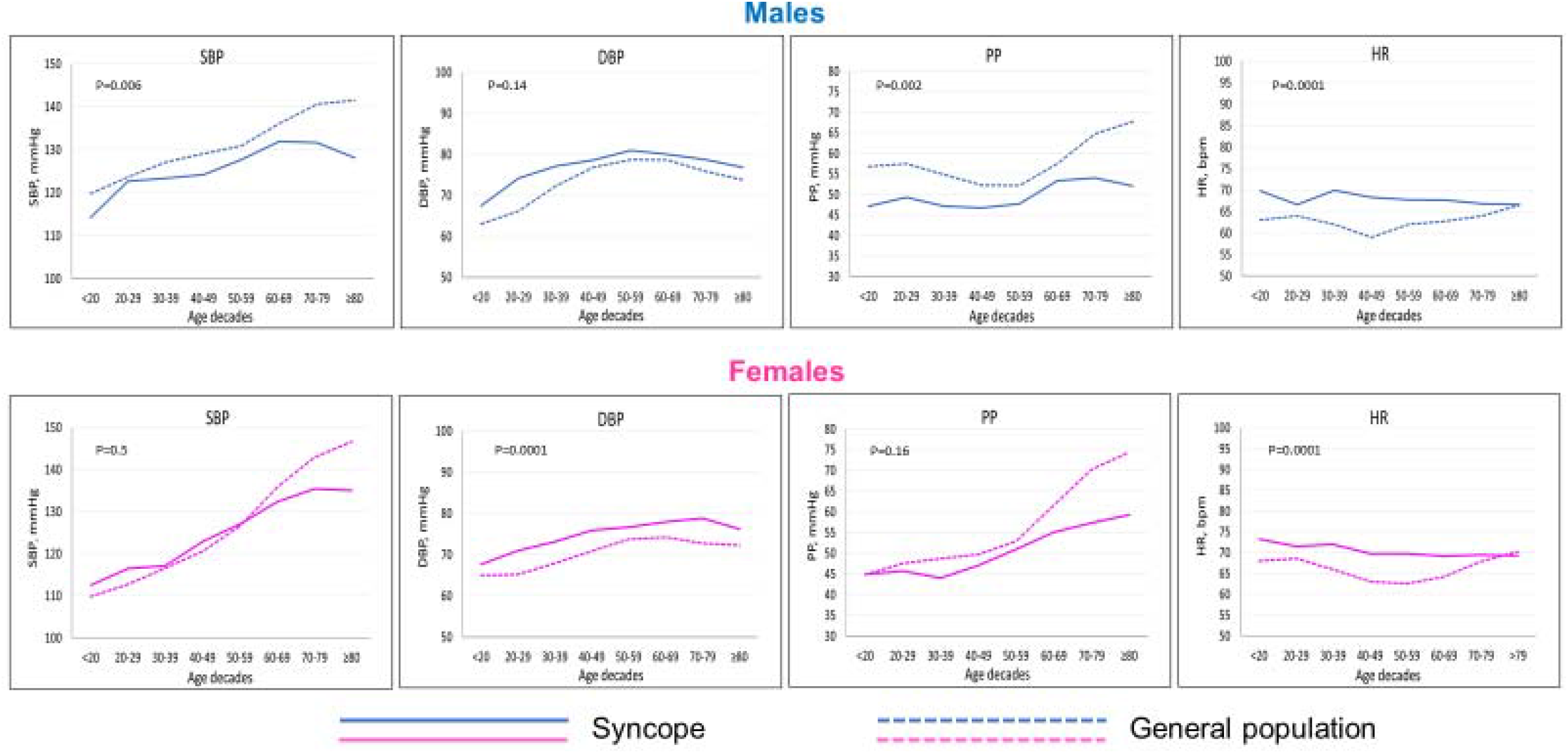
Different hemodynamic profile of systolic blood pressure (SBP), diastolic blood pressure (DBP), pulse pressure (PP) and heart rate (HR) in beats per minute (bpm) in males and females with reflex syncope and in the general population. Data are shown over the different decades, throughout the lifespan.

A sensitivity analysis limited to the 3 cross-sectional studies (intervention trials excluded) showed similar results (*eTable 3 and eFigure 2 and 3*).

The comparison between males and females in the syncope group showed a different pattern of incremental SBP elevation from childhood through older age. **Figure 2** displays changes in SBP levels by decades in ascending order. Compared with males, females had lower SBP at very young age (<20 years) and exhibited a slower increase until the age of 40 and then a steeper increase that continued throughout the rest of life. Therefore, females had lower BP values than males until the age of 40 years and higher values above the age of 60 years. This different pattern is correlated with the male/female ratio calculated at each decade, with males constituting one third of the syncope population until the age of 40 years and approximately one half after that age. The patterns of incremental SBP elevation in the general population, stratified by sex, and their comparisons with the syncope group, are shown in the *eFigure 3*

**Figure 2.**
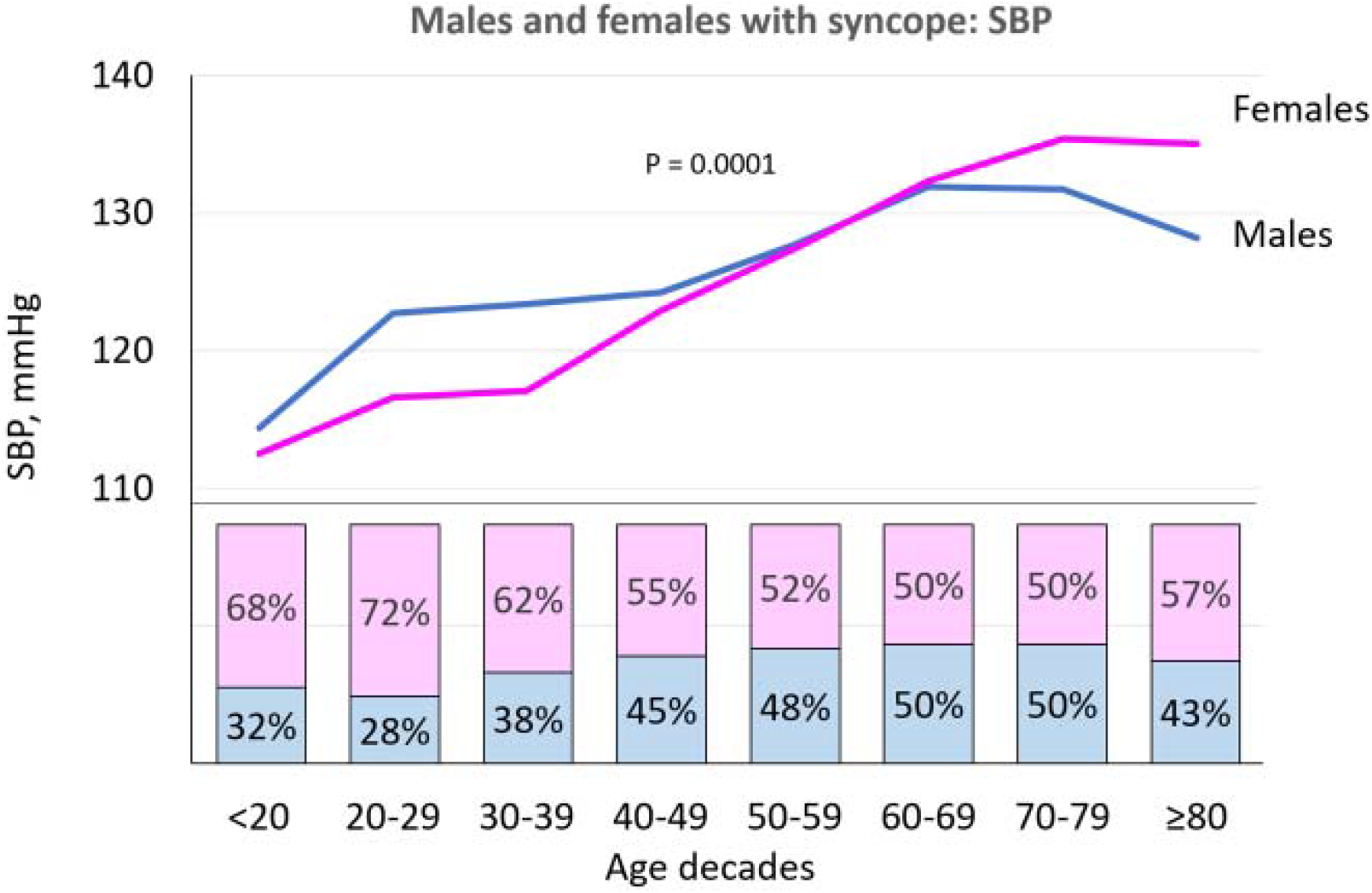
Comparison between gender in patients with reflex syncope. The **upper part** of the figure shows the patterns of incremental systolic blood pressure (SBP) elevation from youth in males and females. Data are shown over the different decades, throughout the lifespan. SBP increases progressively with age. Compared with males, females exhibit a slower increase until the age of 40 and then a steeper increase that continues throughout the rest of life. SBP declines in males after the age of 60 years. A significant interaction of gender with age is present (p=0.0001). The **lower part** of the figure shows the males/females ratio of reflex syncope per age decade. The relative rate of syncope in males and females is related to their SBP until the 6^th^ decade. The slightly higher rate of females at ages ≥80 years is likely due to the higher census prevalence of females in those ages. Chi-square test for trends: p=0.0001

## Discussion

The data presented here for the first time give evidence that BP and HR in patients with reflex syncope differ from the general population. In this multicohort cross-sectional analysis, male patients diagnosed as having reflex syncope showed lower SBP but higher DBP and higher HR compared with population-derived controls. The cardiovascular physiology was similar in females, but lower SBP became evident only over 60 years. According to systolic and diastolic BP alterations, pulse pressure was consistently reduced over decades of life in individuals prone to reflex syncope.

The pathophysiology of reflex syncope, the most common form of transient loss of consciousness, is not completely understood. Reflex syncope is typically triggered either by orthostatic stress or by other stimuli such as emotional distress, somatic and visceral triggers, carotid sinus baroreflexes and neuroendocrine changes (1). The fact that reflex syncope affects approximately one third or more of the population with an apparently benign long-term prognosis in younger individuals, has been a scientific puzzle. In the early 20^th^ century, Cotton and Lewis (20) studied a group of frequently fainting young soldiers presenting with attacks induced either by blood sampling or by standing. They emphasized the role of sudden vasodilation and vagal overactivity in these benign fainting attacks, thus promoting the term “vasovagal faint”. Later, Weissler et al demonstrated that reduction in central blood volume and cardiac output critically contributed to vasovagal reflex initiation (21). However, only after the introduction of non-invasive continuous haemodynamic monitoring techniques in 1980s, laboratory studies on induced vasovagal reflex brought important insights into the pathophysiology of one form of reflex syncope, namely orthostatic vasovagal syncope (VVS). Careful analysis of haemodynamic changes preceding VVS performed independently by different groups of scientists led to the conclusion that orthostatic VVS follows a similar pattern: early stabilization by means of increased HR (which increases cardiac output) and increased arterial vasoconstriction (periphery vascular resistance) followed by circulatory instability, reduced stroke volume and vasovagal reflex activation (22). During the vasovagal reflex, the blood is predominantly pooled in the splanchnic capacitance vessels, which, when the patient is placed in supine position, empty into the right heart through the inferior vena cava and the normal circulation is restored (22). Neuroendocrine studies performed recently revealed that the phase of pre-syncopal circulatory instability is accompanied by a distinct epinephrine and vasopressin surge (23, 24). Hypothetically, this pronounced neuroendocrine response to orthostatic challenge could be evoked by haemodynamic instability and would be secondary to reduced cardiac filling and hypotensive tendency detected by central and arterial baroreceptors. Thus, low SBP would be an important factor facilitating initiation of the neuroendocrine cascade and vasovagal reflex, implying that the different haemodynamics existing in those prone to syncope underpin its occurrence.

In this study, we hypothesized that the cardiovascular profile of patients prone to reflex syncope is primarily characterized by reduced venous return and stroke volume, as evidenced by lower SBP and PP and by compensatory increase in HR and peripheral vascular resistance, the latter represented by elevated DBP **(Figure 3)**. Assuming that aortic compliance in reflex syncope patients is not different from age- and sex-matched general population, PP can be seen as an approximate measure of stroke volume. Therefore, it seems that the patients with reflex syncope have a chronic counter-regulation against reduced stroke volume and low BP (the so called “low BP phenotype”), or that the body has an a priori reduced compensatory capacity leading to intolerance of orthostatic stress challenge (24). The increase in HR and DBP in females below the age of 60 is most likely able to compensate fully for the haemodynamic changes on standing and to preserve SBP (“masked” low BP phenotype). Consequently, the cardiovascular homeostasis, aimed at maintaining brain perfusion and evidenced by the lack of difference in MAP between the groups, is achieved at the expense of chronic or repeated compensatory mechanisms. Reflex syncope manifests when orthostatic stress or other typical triggers overcome the capacity of these adaptative mechanisms to compensate for lower SBP, leading to a decrease in MAP and a reduced blood supply to the brain.

**Figure 3.**
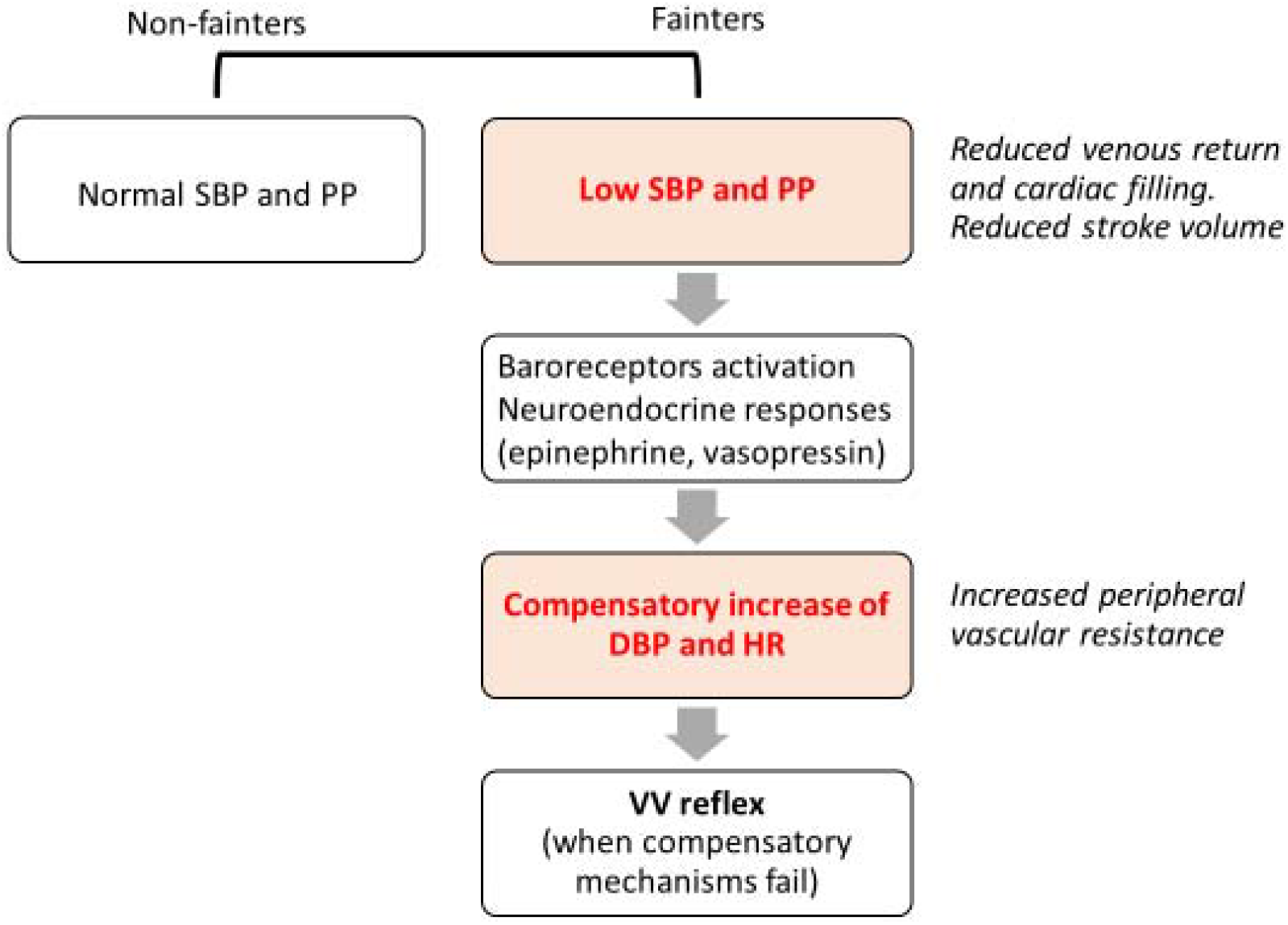
The haemodynamic cascade of orthostatic vasovagal syncope (see text for explanation) SBP=systolic blood pressure; DBP=diastolic blood pressure; HR=heart rate; VV=vasovagal.

There are various possible explanations for this specific cardiovascular profile among patients with reflex syncope. First, these individuals may have lower circulating blood volume or tendency to blood pooling in splanchnic and other subdiaphragmatic regions (4, 24, 25). Women, having lower blood volume in general and less skeletal mass, had on average higher HR, supporting the lower blood volume-hypothesis. Second, the baroreceptors may be chronically reset to a lower mean arterial pressure in syncope patients. It has been demonstrated that baroreceptor sensitivity is elevated in reflex syncope patients, which could explain changes in sympathetic ouflow and elevated DBP and HR, as a consequence (26). Third, the neuroendocrine phenotype, including baseline neuroendocrine levels and their clearance, as well as cardiovascular receptor density and reactivity, may promote lower BP (2-4). Further, supressed chronotropic response and reduction in HR difference between syncope and control group in advanced age might explain the increase in relative SBP difference between the groups as shown in **Figure 1**. A role for hereditary factors seems likely but, yet, they have not been sufficiently explored.

In this study, some 25-30% of all included individuals declared taking antihypertensive drugs. It may be argued that antihypertensive treatment, which is common in later life, may have contributed to the distinctly lower SBP observed among patients older than 60 years. As antihypertensive treatment was balanced between syncope and general population cohorts without potential confounding effect (*eFigure 2)*, the possible impact of higher antihypertensive drug use on lower SBP in the syncope population is unlikely. Alternatively, patients prone to reflex syncope may be more sensitive to antihypertensive medication, which may result in greater BP-reducing effect and lower SBP in this group.

The different lifetime BP profile observed in patients with reflex syncope and in the general population, in both men and in women offer additional insights. Syncope patients showed a lower than expected age-related BP increase, in particular for men >60 years, and women >40 years. Further, women exhibited lower absolute values and less increment in SBP than men until the age of 40 years. Here, the hormonal changes during puberty in females, promoting vasodilation, excessive splanchnic pooling and decreased plasma volume may offer additional explanations (27). The lower SBP might explain why women are more susceptible to syncope than men at younger age **(Figure 2)**. Regardless of intergender differences, both men and women with syncope differ from the general population by higher HR and DBP in youth, which may paradoxically enhance blood pooling by reducing sympathetic muscle activity, whereas SBP curves diverge first at the age of 60, especially in women. An increasing reflex susceptibility among post-menopausal women may plausibly be linked to the lower SBP, in addition to higher HR and reduced pulse pressure observed when younger. Moreover, important anatomical differences between sexes cannot be ignored: women compared with men have not only smaller total body size but also smaller organs, including the heart and the vascular beds; gut and adipose tissue have different susceptibility to pooling and their relative amounts differ between men and women(27).

In men with syncope and in women >60 years old, SBP is consistently lower, PP smaller and HR higher across all age categories, but less in the middle decades. Consequently, the low SBP phenotype seems to be the main determinant of syncope in men and post-menopausal women, while hyperadrenergic activation, expressed by higher HR and elevated DBP component, seem to prevail in younger women. It is well known from epidemiological studies (1) that the incidence of syncope is bimodal, showing a high prevalence in patients aged 10-30, being less common in adults and peaking again after the age of 60 years. Our results might offer an explanation for this pattern. Syncope is more frequent when SBP has a low absolute value or is lower than in the general population. Conversely, syncope is less frequent when SBP of syncopal patients is in the same range as that of the general population **(Figure 2)**. Since this study was a proof-of-concept, no therapeutic SBP targets could be established but our results should prompt further efforts to define the appropriate BP levels for reflex syncope patients in different decades.

These observations may have therapeutic implications in patient selection and provides background for the design of future interventional trials. It is generally known that pharmacological interventions against recurrent reflex syncope are of limited efficacy and randomized studies have not been able to offer a universal treatment outperforming placebo (1). Based on our findings, the therapy should be selected considering the patients’ haemodynamic profile. Younger patients with low-BP phenotype, especially those with extremely low values, might potentially benefit from volume expansion and BP-elevating drugs. In older and hypertensive patients, reduction of antihypertensive agents may prove effective as the patients may move from the “vasovagal reflex corridor” up to the safe BP zone (28). Intensive BP lowering may increase the risk of iatrogenic syncope. Indeed, in the SPRINT study (29) randomization to intensive systolic BP control was associated with a greater risk of adverse events involving hypotension and syncope. During the 5 years of observation, in the intensive BP treatment arm, there were 5.2% patients with primary outcome events (defined as composite of myocardial infarction or acute coronary syndromes, stroke, heart failure, or death from cardiovascular causes) and 3.5% patients with syncope (defined as emergency department visit or serious adverse event) and 3.4% patients with hypotension. Moreover, syncope in older adults, regardless of aetiology, might not be a benign event but rather an indicator of frailty associated with increased cardiovascular risk and mortality (30).

### Limitations

Certain limitations merit consideration. Differences in the years of recruitment of subjects, differences in methods for assessing BP, heterogeneity existing between and within cohort study settings are reported in the *eTable 1*. Information regarding associated risk factors, e.g., body mass index, is lacking. Interpretation of pulse pressure should be made with caution because PP is affected by both stroke volume and vascular properties. In absence of data, we assumed that aortic compliance in reflex syncope patients is not different from age- and sex-matched general population. However, even if a bias due to the above limitation in sampling of patients versus the control group cannot be excluded with certainty, this is unlikely because of the very large population and broad age range of both patients (randomly sampled by referral) and of the general population. Uncertainty exists in any study. Such uncertainty suggests that our results should be confirmed by others.

In order to increase the generalizability of results, we performed a sensitivity analysis, in which we included in the syncope group the results from the 3 cross-sectional databases while excluding the data derived from the 3 clinical trials (*eTable* 3 *and eFigure 2 and 3*).

A similar percentage of subjects in both groups were taking antihypertensive drugs *(eTable 1)*. Lacking individual data of many databases, we were unable to determine the type and dosage used. The interaction between age, prevalence of hypertension and study groups was assessed by a sensitivity analysis *(eFigure 1)* that showed a similar effect of hypertension on syncope group and general population. We have already discussed above the fact that we considered antihypertensive therapy as one of the factors determining the low BP phenotype.

We were unable to determine the proportion of individuals in the control group who had a history of syncope. From the epidemiological studies we can estimate the prevalence of syncope to be approximately 25-30% in the general population, and in most of the cases due to reflex cause.(1) Hypothetically, if these individuals were excluded, the difference in BP and HR between syncope group and control population would, most likely, be even greater. Our syncope patient sample consists of the most symptomatic individuals, who represent 5-10% of the total syncope population. (1) Thus, the corresponding subset of syncope patients in the general population would be around 1-5%.

## Conclusions

The different life-course of blood pressure and heart rate profile that we observed in patients with reflex syncope and in the general population give evidence for a specific cardiovascular profile, lower systolic blood pressure and pulse pressure with higher heart rate and diastolic blood pressure, in patients with reflex syncope which predisposes such subjects to vasovagal reflex. This finding was completely unknown before our study and contributes to our understanding of the mechanism of reflex syncope, i.e., the primary role of cardiovascular haemodynamic differences in the syncope population underpins the genesis of reflex syncope instead of being a readily dismissed neurological phenomenon. The result of this study help to understand why some subjects with similar demographic characteristics develop reflex syncope and others not.

## Data Availability

Data base will be available upon reasonable request to

## Disclouseres

None

