## Supplementary data for "Low blood pressure phenotype underpins the tendency to reflex syncope"

### Supplementary material online

Statistical analysis:

- Generalized Estimating Equation (GEE) models
- Potential effect of hypertension prevalence as a confounder

Sensitivity analyses:

- Supplementary Table S1. Period of recruitment and methodology used for blood pressure measurements in syncope patients and general population
- Supplementary Table S2. Characteristics of syncope study group
- Supplementary table S3. Sensitivity analysis of supine BP and HR limited to the 3 cross-sectional consecutive series of males and females with reflex syncope: comparison with general population
- Supplementary Figure S1. Assessment of the age effect on prevalence of hypertension in syncope group and general population
- Supplementary Figure S2. Sensitivity analysis of supine BP and HR limited to the 3 cross-sectional consecutive series of males and females with reflex syncope: comparison with general population
- Supplementary Figure S3. Comparison between gender and groups in syncope group and general population
- Supplementary References

### Statistical analysis

#### Generalized Estimating Equation (GEE) models

In order to evaluate the relationship of population type (general population and syncope), gender, age decade with BP and HR we employed the Generalized Estimating Equation (GEE) models which take into account the clustering effect due to correlation among estimates within a study. Moreover, we weighted each available mean estimate for the corresponding sample size. The model included four groups of patients according to the gender and population type (syncope or general population): i) male and syncope (MSyn); ii) male and general population (MGP); iii) female and syncope (FSyn); iv) female and general population (FGP). Age decade was included as a continuous variable. To take into account the potential modifying effect of age on the association among groups and BP/HR levels, we included an interaction term between age and groups. The group-specific least square means (and their 95% confidence interval [95% CI]), adjusted for age decade were estimated. The following contrasts of least square means were performed: i)  MSyn vs MGP, ii) FSyn vs FGP to underline differences among population types in males and females. Moreover, we considered a further 3 contrasts; the first two (separately for gender) related to differences in trend of BP/HR levels through age decades among syncope and general population groups and the third related only to syncope patients for differences in trend of BP/HR levels through age decades among males and females.

#### Potential effect of hypertension prevalence as a confounder

We evaluated the potential effect of hypertension prevalence as a confounder veryfing if the relationship between age and prevalence of hypertension changes in function of population type. We modelled the prevalence of hypertension, available only at study level, as a function of average age and population type as well as their interaction

#### Sensitivity analyses

A sensitivity analysis was performed considering only the 3 cross-sectional consecutive series (intervention trials excluded), in order to verify the independence of findings from the potential selection biases of trials

**Table S1.** Main characteristics of included studies and methodology used for blood pressure measurements in syncope patients and general population

| | Years of recruitment | Number of patients | Mean age (years) | Females (%) | Rx for hypertension (%) | Type of study | BP measure | Position and condition |
|---|---|---|---|---|---|---|---|---|
| **Reflex syncope** | | | | | | | | |
| Pool POST trials (a) | 1998-2018 | 664 | 49±24 | 68 | 4 | Trial | Arm cuff manual (POST 1,2,4,5) Finger volume clamp method calibrated with intermittent brachial cuff BP measurements (POST 6) | Post 1-5: sitting Post 6: supine |
| SYSTEMA database (b) | 2008-2018 | 16373 | 50±20 | 74 | 25 | Cross-sectional consecutive series of patients referred for syncope evaluation | Nexfin/Finapress, reconstructed brachial pressure | Supine after 10 min rest, 1 measure average of 30 s period |
| EGSYS trial (c) | 2004 | 136 | 57±24 | 55 | 29 | Trial | Arm cuff manual | Supine after 5 min rest, 1 measure |
| Tigullio database (d) | 2005-2019 | 2818 | 63±20 | 49 | 29 | Cross-sectional consecutive series of patients referred for syncope evaluation | Arm cuff oscillometric automated device | Supine after 5 min rest, 1 measure |
| ISSUE 3 trial (e) | 2007-2011 | 465 | 65±12 | 55 | 32 | Trial | Arm cuff manual | Supine (rest time unknown) |
| Careggi database (f) | 2003-2014 | 806 | 68±18 | 57 | 56 | Cross-sectional consecutive series of patients referred for syncope evaluation | Arm cuff oscillometric automated device | Supine after 5 min rest |
| **General population** | | | | | | | | |
| ALSPAC (g) | 2007 and 2014 | 6946 | 21±6 | 58 | 1 | Population-based | Automated oscillometric device Omron (705 ITfor 17-year cohort and M6 for 24-year cohort | Supine and sitting, after 5 min rest, average of the second and third consecutive readings |
| Malmo Offspring (h) | 2013-2017 | 2506 | 40±14 | 52 | 18 | Population-based | Automated oscillometric Omron M6 device | Average of two measurements in supine position after 5 min rest |
| HUNT 3 (i) | 2006-2008 | 43899 | 53±16 | 55 | 27 | Population-based | Arm cuff automated oscillometric Dynamap | Sitting, after 5 min rest, 2 measures at 1 min interval |
| SCAPIS Malmo (j) | 2014-2018 | 6026 | 57±4 | 53 | 29 | Population-based | Automated oscillometric Omron M6 device | Average of two measurements in supine position after 5 min rest |
| TILDA (k) | 2009-2011 | 4917 | 62±8 | 54 | 33 | Population-based | Finometer | Supine after 10 min rest |

| | | | | | | | | |
|---|---|---|---|---|---|---|---|---|
| ICARe (l) | 1999 | 674 | 74±7 | 59 | 38 | Population-based control group (healthy controls) | Arm cuff manual | Supine after 10 min rest, average of the second and third consecutive readings |

Source:

a) Unpublished pooled data of POST 1,2,4,5,6 trials (ref 1,2,3,4,5), provided by R. Sheldon. Post 5 is not yet published
b) Syncope Study of Unselected Population in Malmö (SYSTEMA), Skåne University Hospital, Malmö, Sweden; unpublished data provided by A. Fedorowski. (ref 6).
c) Unpublished data of EGSYS trial (ref 7), provided by A. Ungar
d) Syncope Unit database at Ospedali del Tigullio, Lavagna, Italy 2006-2019, unpublished data provided by M. Brignole
e) Unpublished data of ISSUE 3 trial (ref 8), provided by M. Brignole
f) Syncope Unit database at Careggi Hospital, Florence, Italy 2003-2014, unpublished data provided by A. Ungar
g) Unpublished data of The Avon Longitudinal Study of Parents and Children (ALSPAC) provided by C Park (ref 9)
h) Unpublished data of the Malmö Offspring Study (MOS), Lund University, Malmö, Sweden provided by Viktor Hamrefors (ref 10)
i) Data available in ref 11
j) Unpublished data of The Swedish CArdioPulmonary BioImage Study (SCAPIS), Lund University and Skåne University Hospital, Malmö, Sweden Viktor Hamrefors (ref 12)
k) Unpublished data of TILDA study (ref 13) provided by RA. Kenny
l) Unpublished data of the ICARe study (ref 14) provided by A.Ungar

**Table S2**. Characteristics of syncope study group

| | Diagnosis | History of syncope | Cardiac/ECG abnormalities |
|---|---|---|---|
| **Reflex syncope** | | | |
| Pool POST trials (a) | Clinical (Calgary syncope score) | Post 1: 9 (IQR 5-20)<br>Post 2: 15 (2.3 per year)<br>Post 4: n.a.<br>Post 5: n.a..<br>Post 6: 11 (4-20) | Post 1:4%<br>Post 2: absent<br>Post 4:absent<br>Post 5: absent<br>Post 6: absent |
| SYSTEMA database (b) | Clinical (tilt pos and tilt neg) | 1 episode: 12.5% (n=207)<br>>1 episode: 87.5%<br>Median: 5 (IQR 2-10) | EF <55%<br>21.6% |
| EGSYS trial (c) | Clinical + web interactive tool | 1 episode: 42%<br>>1 episode: 58% | 19% |
| Tigullio database (d) | Clinical (tilt pos and tilt neg) | 1 episode: 952 (34%)<br>>1 episode: 1846 (66%) | 418 (15%) |
| ISSUE 3 trial (e) | Clinical (tilt optional) | Median 7 (IQR 4-10) | 12% |
| Careggi database (f) | Clinical (tilt pos and tilt neg) | Median 2 (IQR 1-5) | 26% |

Source:

a) Unpublished pooled data of POST 1,2,4,5,6 trials (ref 1,2,3,4,5), provided by R. Sheldon. Post 5 is not yet published
b) Syncope Study of Unselected Population in Malmö (SYSTEMA), Skåne University Hospital, Malmö, Sweden; unpublished data provided by A. Fedorowski. (ref 6).
c) Unpublished data of EGSYS trial (ref 7), provided by A. Ungar
d) Syncope Unit database at Ospedali del Tigullio, Lavagna, Italy 2006-2019, unpublished data provided by M. Brignole
e) Unpublished data of ISSUE 3 trial (ref 8), provided by M. Brignole
f) Syncope Unit database at Careggi Hospital, Florence, Italy 2003-2014, unpublished data provided by A. Ungar

**Table S3.** Sensitivity analysis of supine BP and HR limited to the 3 cross-sectional consecutive series of males and females with reflex syncope: comparison with general population

| | Syncope | | | | General population | | | | P value |
|---|---|---|---|---|---|---|---|---|---|
| | N° of pts | Observed mean | LS adjusted mean | 95% CI | N° of pts | Observed mean | LS adjusted mean | 95% CI | |
| **Males** | | | | | | | | | |
| Systolic blood pressure, mmHg | 2365 | 127.9 | 126.3 | 128.9 - 133.8 | 29424 | 131.2 | 131.4 | 128.9 - 133.4 | 0.0001 |
| Diastolic blood pressure, mmHg | 2365 | 78.1 | 77.4 | 75.0 - 79.8 | 29424 | 75.1 | 75.3 | 73.9 - 76.6 | 0.14 |
| Pulse pressure, mmHg | 2365 | 48,8 | 49.0 | 44.8 - 53.2 | 29424 | 57.9 | 56.2 | 54.1 - 58.2 | 0.003 |
| Mean arterial pressure, mmHg | 2365 | 93.0 | 93.7 | 91.9 - 95.4 | 29424 | 92.5 | 93.7 | 92.4 - 95.5 | 0.81 |
| Heart rate, bpm | 2019 | 67.3 | 67.9 | 66.4 - 69.3 | 9493 | 67.3 | 62.7 | 60.4 - 64.9 | 0.0001 |
| **Females** | | | | | | | | | |
| Systolic blood pressure, mmHg | *2886* | 128.2 | 126.0 | 121.6 - 130.5 | *35554* | 125.7 | 126.8 | 124.5 -129.2 | 0.83 |
| Diastolic blood pressure, mmHg | *2886* | 74.0 | 74.8 | 73.9 - 75.8 | *35544* | 71.0 | 71.3 | 70.3 - 72.4 | 0.001 |
| Pulse pressure, mmHg | *2886* | 51.1 | 51.2 | 46.1 - 56.3 | *35544* | 56,2 | 55.5 | 52.2 - 58.8 | 0.16 |
| Mean arterial pressure, mmHg | *2886* | 91.6 | 91.9 | 90.6 - 93.1 | *35544* | 89.0 | 89.8 | 89.4 - 90.2 | 0.002 |
| Heart rate, bpm | *2426* | 70.3 | 70.5 | 69.6 - 71.5 | *11576* | 65.7 | 65.0 | 62.3 - 67.6 | 0.0001 |

LS=least square

**Figure S1.**

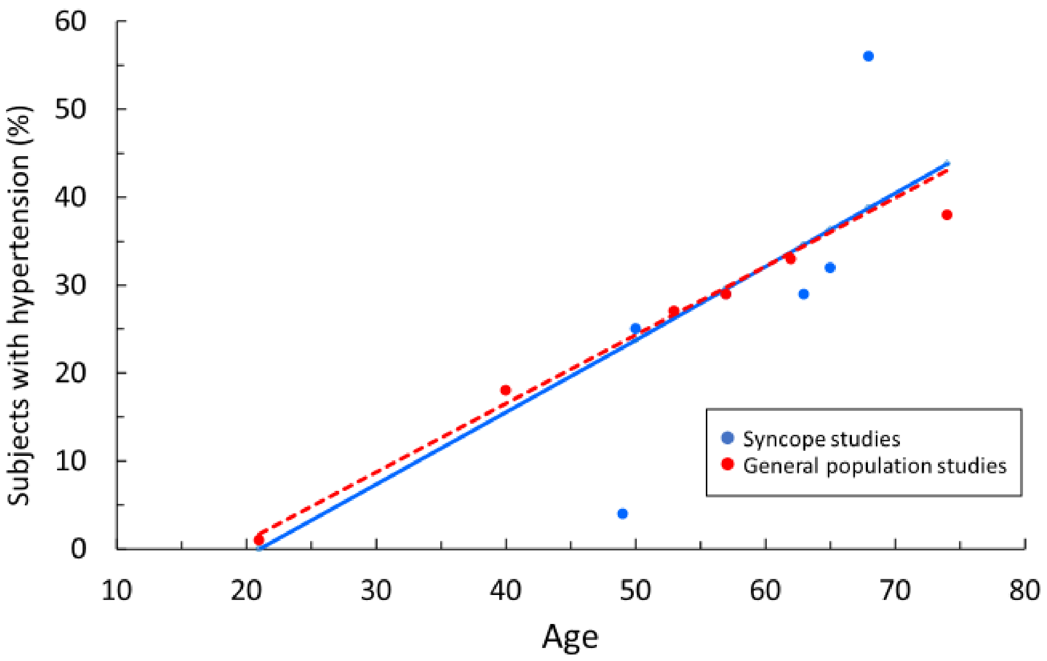


**Figure S1.** Assessment of the age effect on prevalence of hypertension in syncope group (blue bullet and line) and general population (red bullets and line). Antihypertensive therapy was balanced between syncope and general population cohorts without potential confounding effect (p=0.89).

#### Figure S2

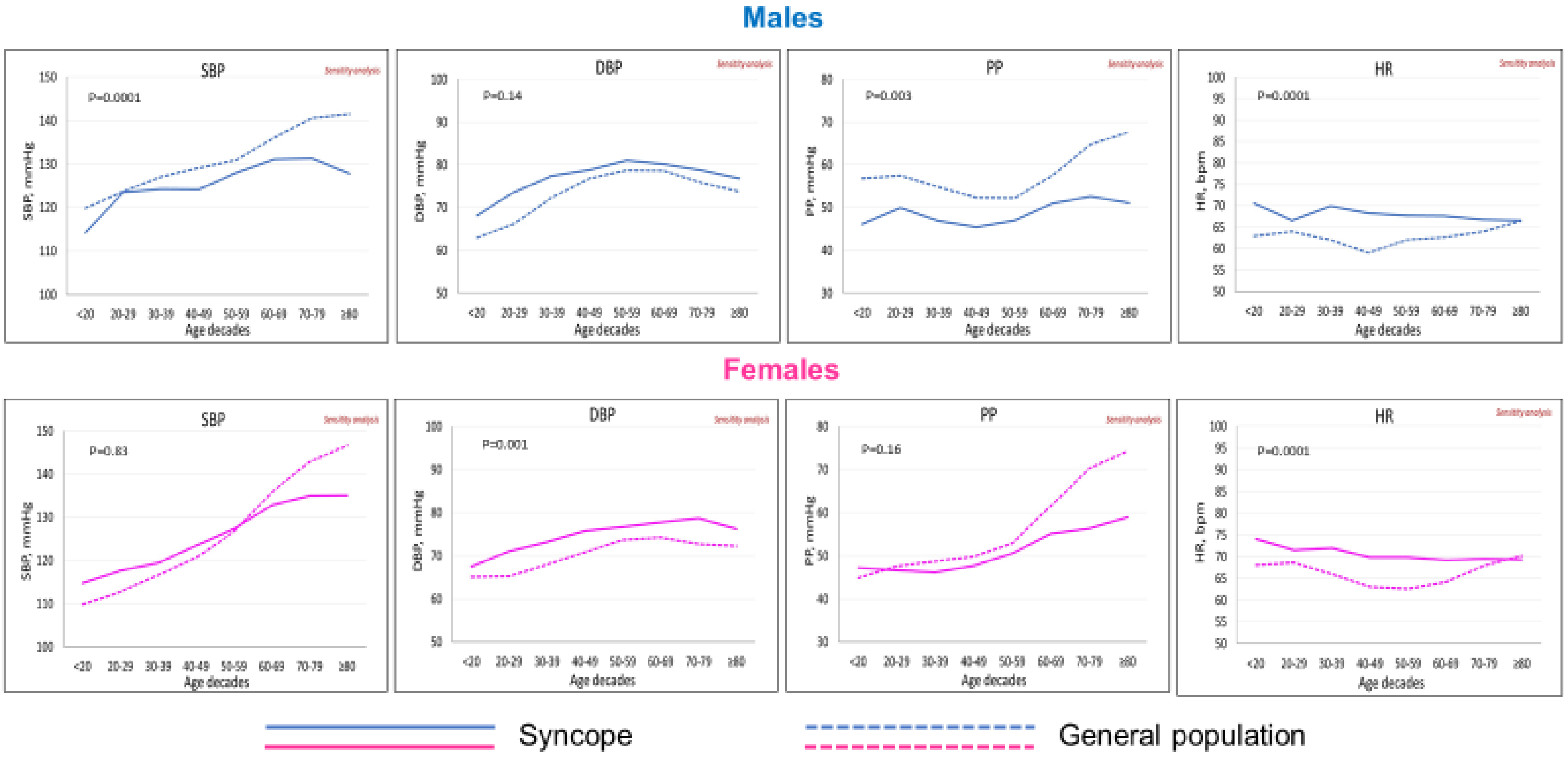


**Figure S2.** Sensitivity analysis limited to the 3 cross-sectional consecutive series of patients with reflex syncope (ref b,d and f of supplementary table 1). Life-course trajectories of systolic blood pressure (SBP), diastolic blood pressure (DBP), pulse pressure (PP) and heart rate (HR) in males and females with reflex syncope and in the general population.

**Figure S3.**

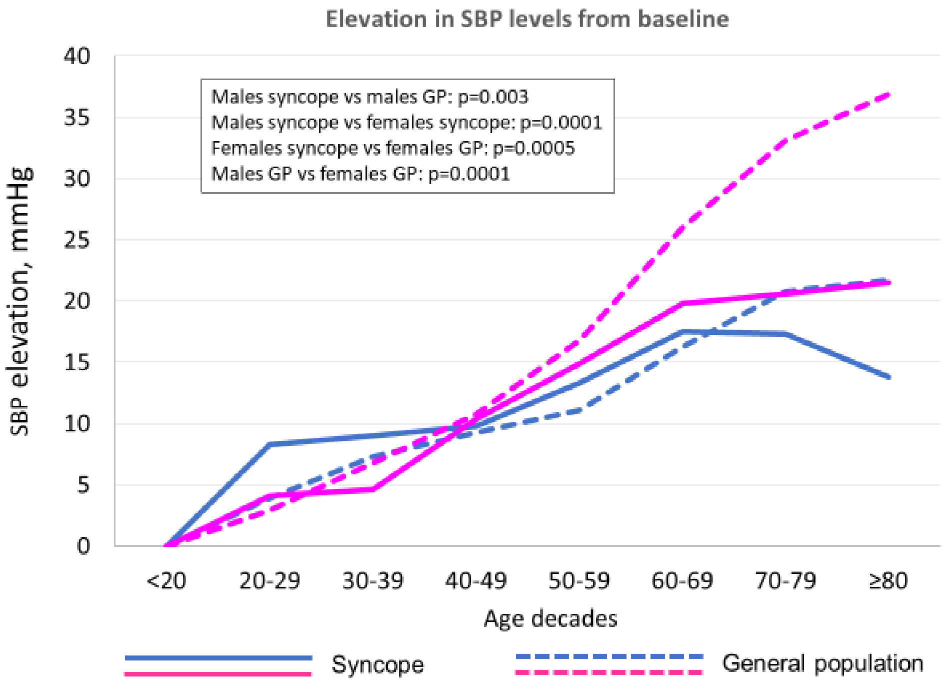


**Figure S3.** Comparison between gender and groups. The figure shows the patterns of incremental systolic blood pressure (SBP) elevation from youth in males and females of syncope and general population groups. Data are shown over the different decades, throughout the lifespan. SBP increases progressively with age. Compared with males, females exhibit a steeper increase from the age of 40 years that continues throughout the rest of life either in syncope group and in general population group. Compared with the syncope group, the general population group exhibit a steeper increase either in males (from the age of 60 years) and in females (from the age of 40 years) that continues throughout the rest of life.
